# A phenotypic spectrum of autism is attributable to the combined effects of rare variants, polygenic risk and sex

**DOI:** 10.1101/2021.03.30.21254657

**Authors:** D Antaki, A Maihofer, M Klein, J Guevara, J Grove, Caitlin Carey, O Hong, MJ Arranz, A Hervas, C Corsello, AR Muotri, LM Iakoucheva, E Courchesne, K Pierce, JG Gleeson, E Robinson, CM Nievergelt, J Sebat

## Abstract

The genetic etiology of autism spectrum disorder (ASD) is multifactorial with contributions from rare variants, polygenic risk, and sex. How combinations of factors determine risk for ASD is unclear. In 11,313 ASD families (N = 37,375 subjects), we investigated the effects rare and polygenic risk individually and in combination. We show that genetic liability for ASD differs by sex, with females having a greater polygenic load, and males having a lower liability threshold as evident by a negative correlation of rare and polygenic risk. Multiple genetic factors were associated with differing sets of behavioral traits with effects that differed by sex. Furthermore, the correlation of parental age with genetic risk for ASD was attributable to *de novo* mutations and sex-biased effects of inherited risk in parents. Our results demonstrate that a phenotypic spectrum of ASD is attributable to the relative loadings and gene-by-sex effects of rare and common variation.

## Intro

The major risk factors for Autism Spectrum Disorder (ASD) are genetic, and these include multiple types of rare and common variation. At one extreme of the genetic architecture of ASD are the rare *de novo* mutations (DNMs), including copy number variants (CNVs) ^1^ and protein-truncating SNPs and indels ^2^, in which a major contributor to risk is a dominant-acting variant. At the other extreme is polygenic risk that is measured as the sum of thousands of common risk alleles with small effects ^3^. Despite the success in identifying and characterizing multiple types of genetic risk, there is no one rare variant, gene or polygenic score that has a high predictive value for an ASD diagnosis. Even copy number variants (CNVs) with large effects sizes for ASD (e.g. OR>30) present with variable psychiatric traits ^4^, and risk is attributable to a combination of rare and common variation ^5,6^.

Sex is also a major genetic factor that influences ASD risk. Males are diagnosed with ASD more frequently than are females at a ratio of 4:1. A small proportion of cases are associated with X-linked variants ^7^, but the male preponderance of ASD is not explained predominantly by genetic variation on sex chromosomes. We and others have hypothesized that it may instead be attributable to sex differences in the effects of autosomal variants ^8-10^. This hypothesis is supported by previous studies showing that females with ASD have a greater burden of rare CNVs ^1,11,12^ and gene mutations ^13,14^. However, the nature of gene-by-sex interactions in ASD has not been analyzed systematically.

Previous genetic studies have been focused on defining new categories of rare variant risk from DNA sequencing or by improving the statistical power of genome wide association studies (GWAS). How combinations of multiple genetic factors contribute to risk and to clinical presentation is not known. Here we investigate, in a large dataset of whole genomes and exomes, the combined contributions of *de novo*, rare inherited and polygenic risk to ASD. We show that the genetic architecture of ASD varies as a spectrum of rare and common variation, each having distinct phenotypic correlates and differential effects in males and females.

### Defining multiple components of genetic risk

We investigated the combined effects of multiple genetic factors, detectable by genome sequencing or a combination of exome and SNP genotyping, on risk for ASD. We focus on several genetic factors that have established associations with case status, such as de novo protein-truncating (dnLoF) and missense (dnMIS) mutations ^1,2^ and rare inherited variants ^15,16^ that disrupt genes (inhLoF) and polygenic scoring models that have been associated with ASD case status including polygenic scores for ASD (PS_ASD_), schizophrenia (PS_SZ_) and educational attainment (PS_EA_) ^17,18^.

We confirmed the associations of genetic factors with ASD by whole genome analysis of 37,375 subjects from 11,313 ASD families (12,270 cases, 5,190 typically-developing siblings and 19,917 parents. The sample was comprised of three datasets, including whole genome sequencing (WGS) of cohorts from UCSD (https://sebatlab.org/reach-project) and the Simons Simplex Collection (SSC) and exomes and SNP genotyping from the SPARK study ^19^ (see methods and **Table S1**). SNP, indel, structural variant (SV), and DNM calling, were performed using functionally equivalent pipelines for each dataset consisting of GATK best practices and our laboratory’s SV calling practices (see online methods). SNP genotypes from WGS and imputed genotypes from GWAS were merged and filtered to strand unambiguous SNPs. Principal components were calculated to correct for population stratification and related confounders. PRS were calculated from summary statistics of current GWAS meta-analyses of autism, educational attainment, and schizophrenia. Variants were annotated for gene functional constraint. Analysis of rare protein-truncating (LoF) and cis-regulatory (CRE) variants were restricted to genes that show significant variant-intolerance (LOEUF>0.37) and analysis of missense variants were restricted to those with missense badness (MPC) scores > 2.

Association tests were performed for case-control differences in DNM burden. Association of inherited risk was tested by transmission disequilbrium test (TDT) ^15^. Similarly, common variant associations were tested by polygenic TDT (pTDT) that measures overtransmission of risk alleles as the deviation in polygenic scores between offspring and the average score of the parents ^17^. Results demonstrate significant contributions from genetic factors including *de novo* loss of function (dnLoF) and missense (dnMIS) mutations (**Fig. 1A**), rare inherited gene disrupting SNVs (inhLoF) and SVs (LoFSV) and SVs that disrupt cis-regulatory variants (CRE-SVs) of constrained genes (**Fig. 1B**); and polygenic scores PS_ASD_, PS_SZ_ and PS_EA_ (**Fig. 1C**).

**Figure 1.**
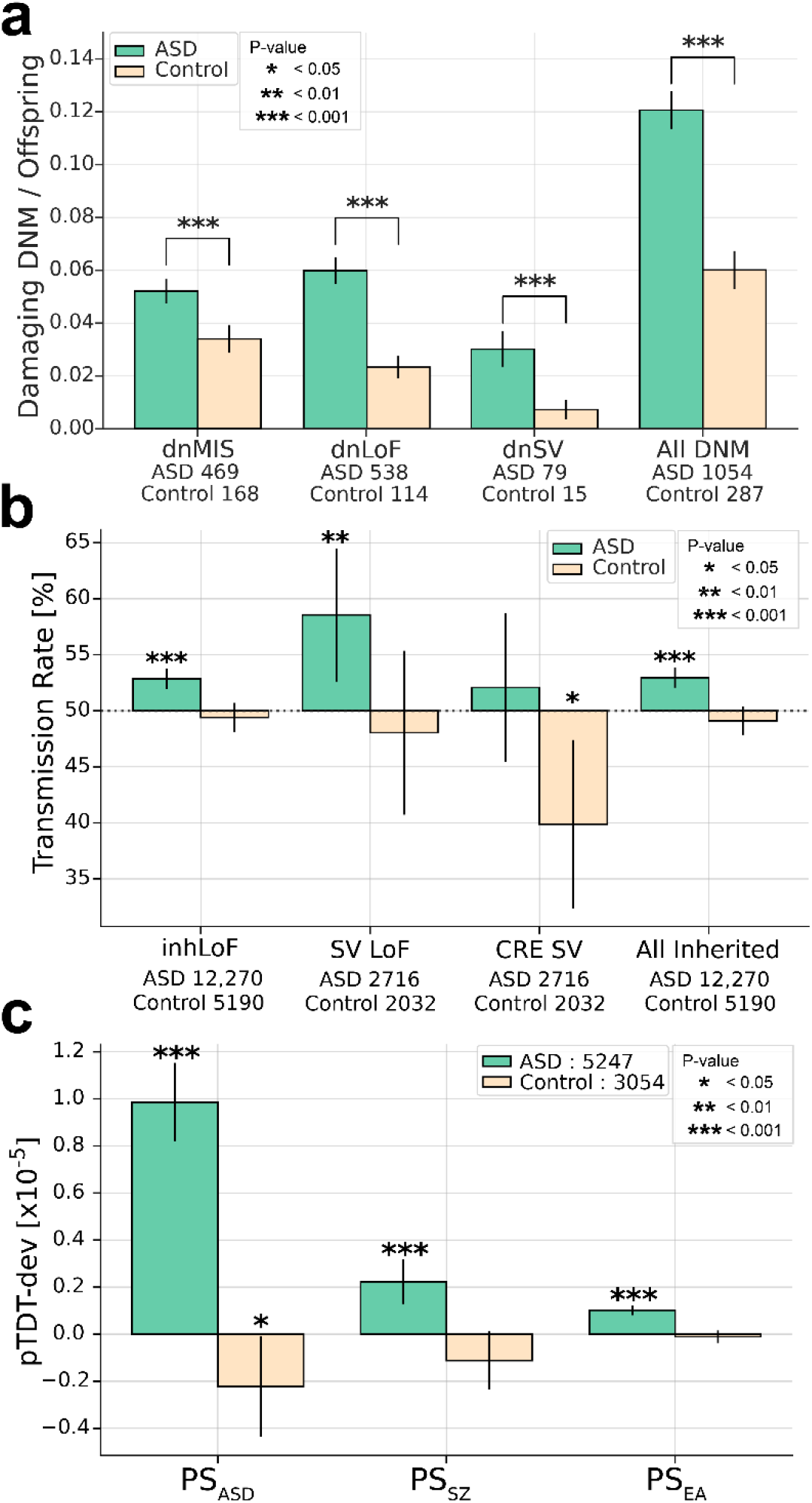
Risk for ASD is attributable to multiple genetic factors including DNMs, rare inherited variants and polygenic risk. Multiple genetic factors that have been previously associated with ASD were confirmed in our combined sample. **(A)** Damaging DNMs in genes that are functionally constrained (LOEUF < 0.37 and MPC ≥ 2), including missense variants (dnMIS), and protein-trunctating SNVs and indels (dnLoF) and SVs (dnSV) occur at higher frequencies in case than in sibling controls (**B**) Protein-truncating SNVs and indels (inhLOF) and SVs (SVLoF) and non-coding SVs that disrupt cis-regulatory elements (CRE-SVs) were associated with ASD based on TDT test. (**C**) Polygenic TDT (pTDT) was significant for all three polygenic scores for autism (PSASD), schizophrenia (PSSZ), and educational attainment (PSEA). Results for panels A, B and C and full lists of rare *de novo* and inherited variants in constrained genes are provided in **supplementary tables 2, 3 and 4** respectively.

We examined the combined effects of rare and common variation in the combined sample. To ensure that rare and common genetic factors are ascertained consistently across the three cohorts, analysis was restricted to six categories that are detectable in exome and WGS with comparable sensitivity: dnLoF, dnMIS, inhLoF and polygenic scores (PS_ASD_, PS_SZ_, PS_EA_). SVs and CNVs, variant types that cannot be ascertained comparably in exome and WGS datasets, were not included. In addition, to minimize ancestry as a confounder in polygenic scores, analysis was restricted to a subset of 7,181 families (N = 25,391 subjects) with parents and offspring that have confirmed European ancestry.

The contribution of each factor individually and the additive contributions of rare variants, polygenic risk and six factors combined was estimated by multivariable regression (**Fig. 2A**). The variance explained by individual genetic factors in our datasets was consistent with previous studies. Polygenic risk explained from 1% to 2.4% of the variance in case status across the three cohorts (**supplementary table 5)** consistent with the ∼2% of variance explained by polygenic risk in the most recent GWAS meta-analysis ^3^. Our results indicate that rare variants and polygenic risk form two major components of the genetic architecture of ASD, and the additive effects of all factors combined could be quantified in a single model (r^2^ = 3.3%, **supplementary table 5**). We applied the estimates of the multivariable regression to create composite genetic risk scores of multiple factors including a rare variant score (RVRS) for the combination of dnMis, dnLoF and inhLoF, a polygenic score (PRS) for the combination of PS_ASD_, PS_SZ_ and PS_EA_, and a genomic risk score (GRS) for the combination of all six genetic factors. For each risk score, we calculated the case-control odds ratios at multiple score thresholds, and we find that across the full distribution of risk scores, the GRS detects an effect size that is >40% stronger than effect sizes for PRS or RVRS (**Fig. 2B, supplementary table 5**)

**Figure 2.**
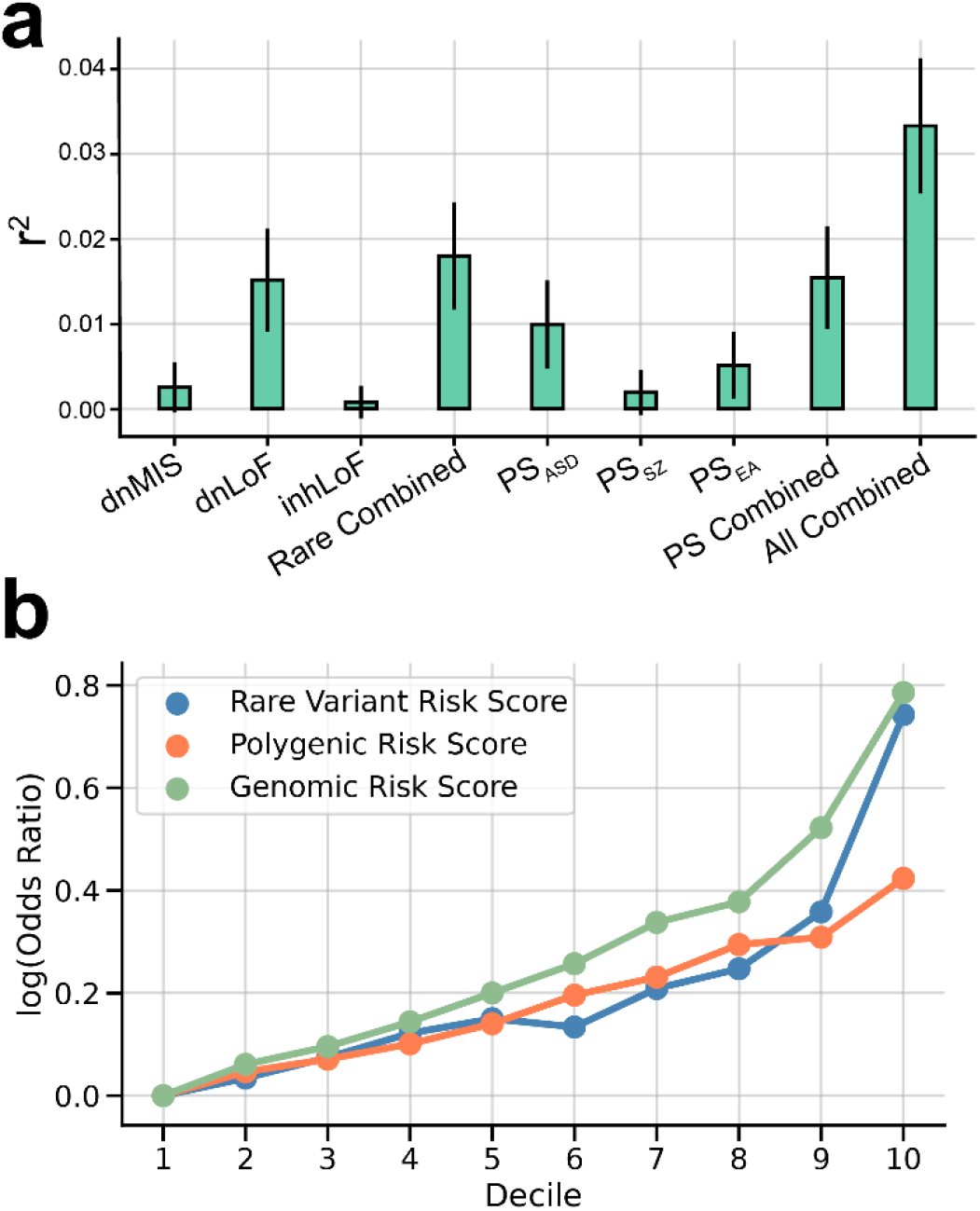
Multivariable regression of 6 genetic factors to create a combined (rare and polygenic) risk score. (**A**) Variance in case status explained (r2 and 95% CI) by each genetic factor individually and in combination. Combined effects of rare variants (Rare combined) polygenic scores (PS combined) and all genetic factors (All combined) were estimated by multivariable logistic regression controlling for sex, cohort and principal components. (**B**) log_10_ odds-ratios of case/control proportions for the composite genetic risk scores RVRS, PRS, and GRS at multiple thresholds (deciles). Across all thresholds, effect sizes for the GRS was 41-42% greater than for RVRS or PRS alone. See results in **supplementary table 5**

### Sex differences in genetic load and genetic liability

The burden of common and rare genetic risk was greater in female cases compared to males (**Fig 3a-b**). Female cases had significantly increased load of combined rare (P = 2e-6, **Fig. 3a**) and combined polygenic scores (P = 2.17 e-04, **Fig. 3b**) compared to male cases, while the genetic loads in controls did not differ significantly by sex. These results are consistent with a “female protective effect” in which a greater genetic load is required for females to meet the diagnostic criteria for ASD ^20^. The full distribution of GRS is skewed upward in females compared to males (**Fig. 3C**), which is further highlighted by a fill plot comparing the densities of distributions of GRS between groups (**Fig. 3D**). As expected, the distribution of GRS is bimodal with a subset of DNM carriers having the highest scores and the greatest enrichment of female cases.

**Figure 3.**
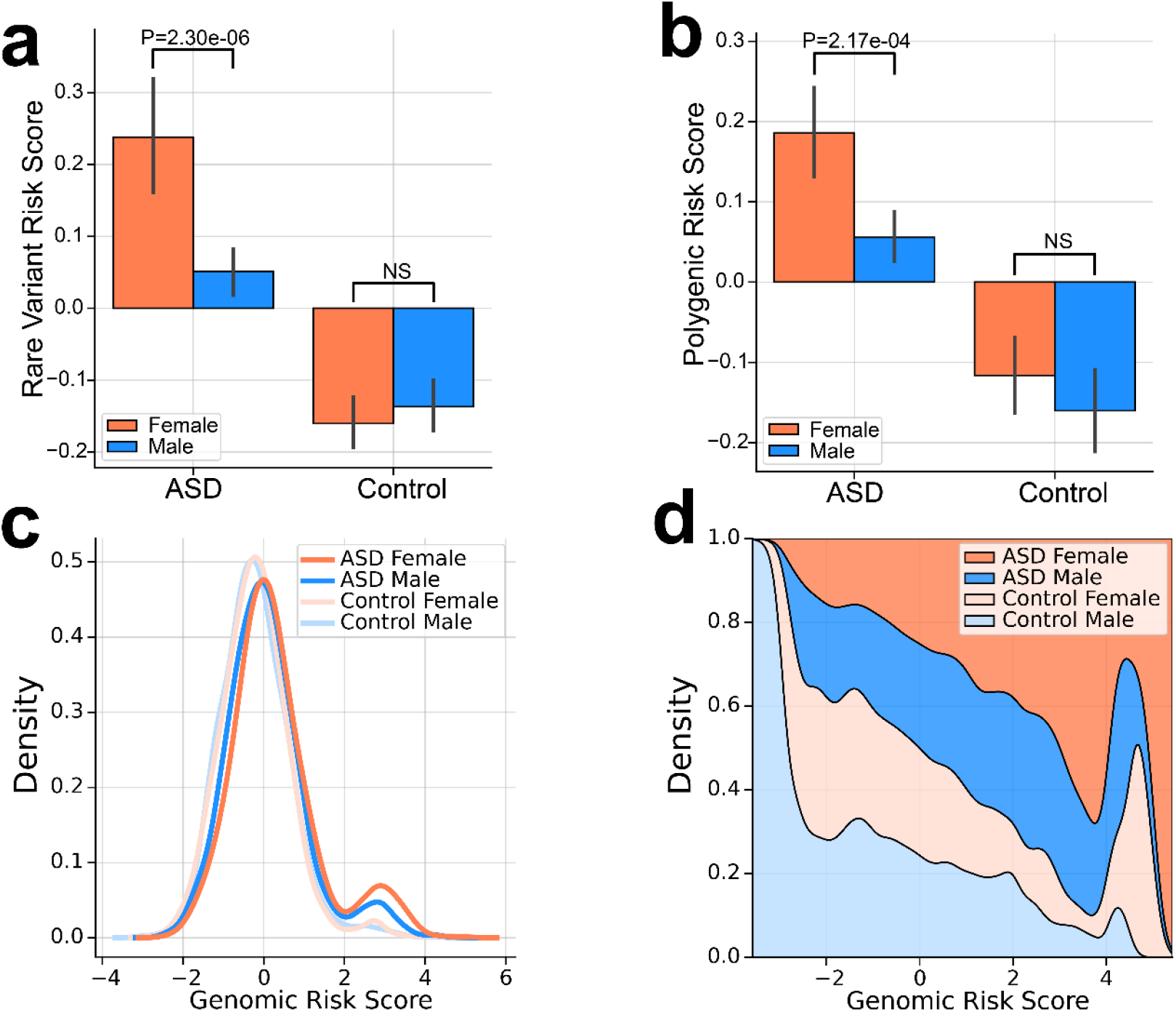
Increased genetic load in females with ASD compared to males. Increased burden of genetic risk in female cases compared to male cases is evident for **(a)** combined rare *de novo* and inherited variants (RVRS) and **(b)** combined polygenic scores (PRS). P-values from a 2-sample t-test are shown. **(c)** Sex differences in the combined genetic load (GRS) is evident across the full distribution. **(d)** A fill plot comparing the densities of distributions illustrates that the GRS of females (cases and controls) are skewed upward relative to males.

According to a liability-threshold model for ASD ^21,22^, a total genetic load sufficient to meet the diagnostic criteria for disease can be reached through differing combinations of rare and common variation. Subjects having a rare variant of large effect may require less polygenic load ^5^ and vice versa. In this study we observed that male cases who carry *de novo* damaging mutations (dnLoF or dnMIS) had a combined polygenic load that was reduced compared to other males with ASD (P = 0.017 **Fig. 4A**). Thus, in the presence of a damaging DNM, less polygenic risk is required to meet diagnostic criteria for ASD. By contrast the combined polygenic load did not differ significantly in female cases with and without DNMs.

**Figure 4.**
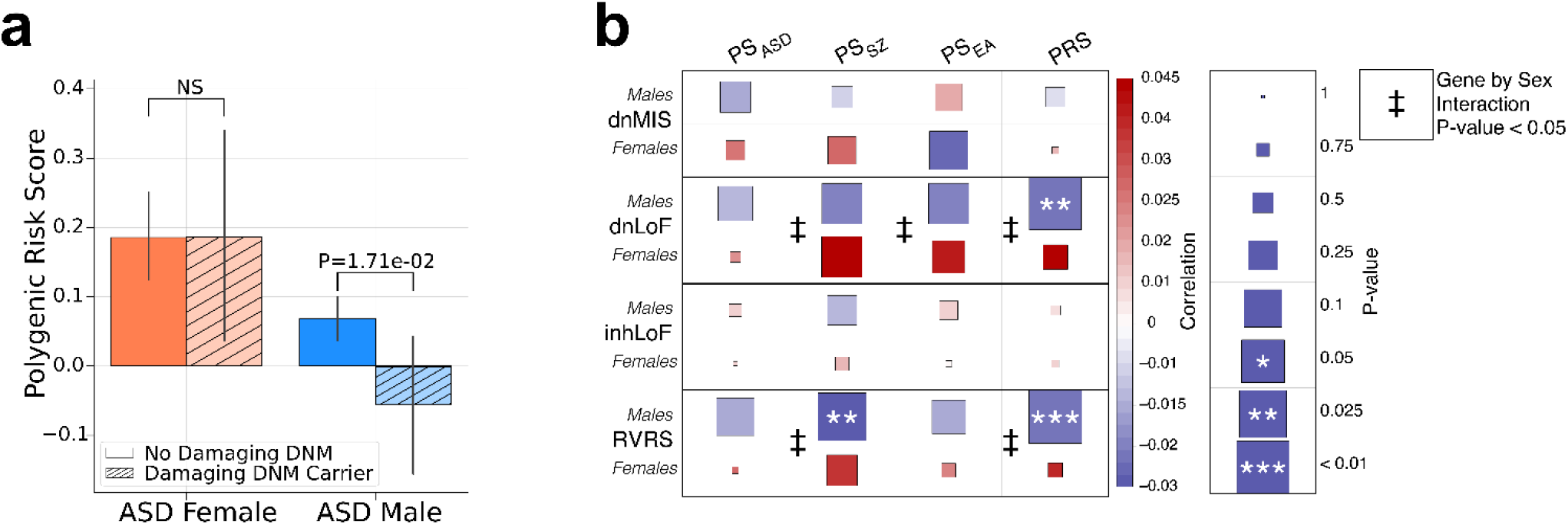
Significant liability threshold for rare variants and polygenic risk in males. **(A)** Transmission of lygenic risk (pTDT) is reduced to males that carry damaging DNMs (dnLoF and dnMIS combined), but not to males. **(B)** A heatmap displaying the strength of the correlations between polygenic scores and rare variants. egative genetic correlations of polygenic scores and rare variants is male-biased, consistent with a liability reshold model. Results are a provided in **supplementary table 6**.

We further investigated sex differences in genetic liability threshold by testing the pairwise correlations of multiple factors by linear regression with sex as an interaction term (**Fig. 4B**). A significant effect of sex was detected for the correlation of DNMs with multiple polygenic scores (PS_EA_, PS_SZ_ and PRS). In all cases, there was a negative correlation of dnLoF with PS in males but not in females. These results provide evidence of a threshold for genetic liability in males in which carriers of DNMs have significantly reduced polygenic risk. A lack of correlation of dnLoFs and PRS in females was consistent with females having a greater threshold for risk and a requirement for greater overall load of both rare and polygenic variants. The above results demonstrated wide variation in the relative loadings of rare and common variation in our ASD cohorts, and the more limited liability threshold in males suggests that the genetic architecture in males may represent a spectrum of genetic loadings from *de novo* to predominantly polygenic. We also see a negative correlation of rare inherited variants (inhLoF) and DNMs (P = 0.03, **Supplementary Fig. 1A**), consistent with a liability threshold model, and we find weak evidence for a sex bias in the transmission of inhLoFs (**Supplementary Fig. 1B**). See supplementary material for additional discussion on the influence of sex on the transmission of rare variants in families.

### Differential effects of rare and common variants on behavioral traits

We hypothesize that the differences in genetic architecture that we observe in this study could underlie variation in clinical phenotype across the autism spectrum. DNMs have been associated with a more severe clinical presentation characterized by greater intellectual impairment ^2,23^ and delays in meeting developmental milestones ^24^.

Likewise, polygenic scores for cognitive traits have stronger effects in high-functioning “Asperger” cases compared to other clinical subtypes of ASD ^18^. However, previous studies have not systematically examined the contributions of rare variants and polygenic risk to symptom severity in ASD families. We investigated behavioral correlates of genetic factors in quantitative phenotype data on cases, sibling controls and parents that was available in the SSC and SPARK cohorts. We quantified the effects of genetic factors on diagnostic measures in offspring including repetitive behavior (RBS), social responsiveness (SRS), social communication (SCQ), adaptive behavior (VABS) and motor coordination (DCDQ). We also investigated genetic effects on behavioral traits in parents, including social impairments (SRS, BAPQ), Educational Attainment (EA), and parental age at birth of the proband. Genetic effects were tested by linear regression controlling for cohort, age, sex and principal components, and effects were also tested for gene-by-sex interactions.

Different genetic risk factors for ASD had distinct patterns of correlated traits. DnMIS and dnLoF were associated with impairments in social communication, motor coordination and adaptive function in cases and were associated with social deficits in sibling controls (**Fig. 5A**). Polygenic scores (PS_EA_ and PS_SZ_) were associated with a range of behavioral traits including repetitive behavior, social communication and adaptive function. The patterns of trait correlation for polygenic scores were surprising in some respects. Despite the significant contribution of PS_ASD_ to case-control status, PS_ASD_ had a modest effect on ASD symptom severity. Variation in clinical features was correlated more strongly with other factors that were, to some degree, orthogonal to case status. Also, surprising, despite its positive correlation with case status, PS_EA_ was negatively correlated with severity of repetitive behavior and deficits in social communication. While it is intuitive that the severity of ASD symptoms was associated with genetic predictors of cognitive impairment (low PS_EA_ and high dnLoF, dnMIS, inhLOF), more work is needed to understand why case status is associated with genetic predictors of high educational attainment. For instance, the association of high PS_EA_ with ASD may be driven by a subgroup of cases that are high-functioning 18 (see supplementary materials for further discussion).

**Figure 5.**
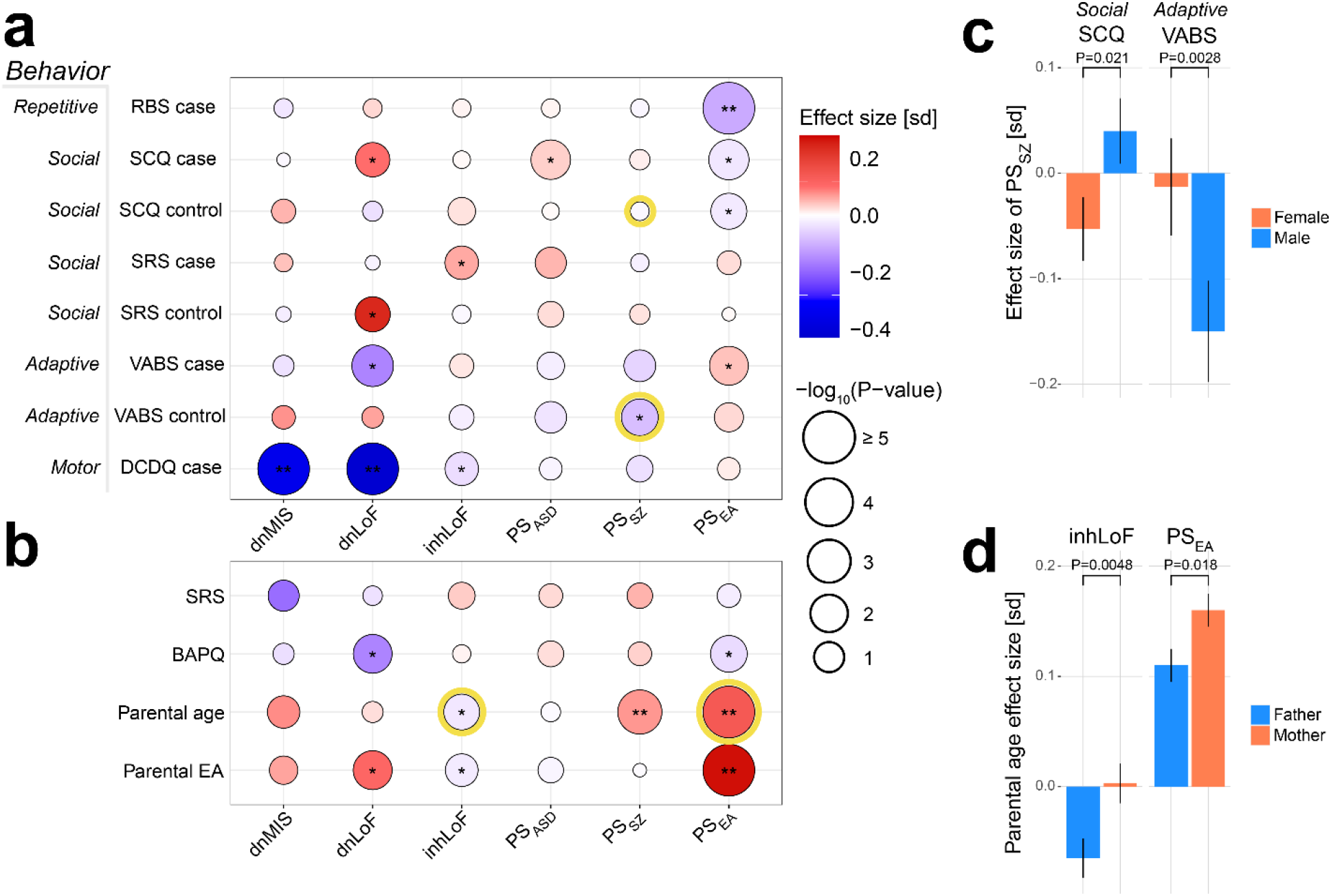
Differential effects of rare and common variation on behavioral traits in cases, sibling controls and parents. **(A)** The effects of genetic factors were tested on five phenotype measures in children: repetitive behavior (RBS), social responsiveness (SRS), social communication (SCQ), vineland adaptive behavior (VABS) and developmental motor coordination (DCDQ). Gene-phenotype correlations were tested by linear regression controlling for sex, age, cohort and PCs. Effect size is given as standard deviation (sd) of phenotype per unit of genetic factor. (**B**) Genetic effects on parental behavior were tested for autism-related symptoms (BAPQ, SRS), educational attainment and parental age. Six gene-trait correlations were significant after bonferroni correction for 72 tests (**P ≤ 0.0007), fourteen were nominally significant (*P ≤ 0.05), and four showed evidence of sex-biased effects (gene*sex interaction P ≤ 0.05 highlighted in yellow. Note, RBS, SRS, SCQ and BAPQ are measures of “deficit”; thus **red** corresponds to increased severity. VABS and DCDQ are measures of “skill”; thus **blue** corresponds to increased severity on these two instruments. **(C)** PSSZ had male-biased effects on social communication and adaptive function in offspring. **(D)** Rare inhLoF variants carried by parents were associated with earlier parental age, specifically in fathers. PSEA was associated with advanced parental age with an effect that was stronger in mothers. Sample sizes for each phenotype ranged from 3,429 to 11,485. Sample numbers and results are summarized in **supplementary table 7**.

In addition to genetic risk having effects on symptom severity in cases and their siblings, genetic risk factors were also associated with behavioral traits in their adult parents. Specifically, DNMs in cases were associated with increased educational attainment and reduced ASD symptoms (BAPQ) in parents, consistent with a *de novo* etiology (**Fig. 5B**). Similarly, polygenic scores for educational attainment (PS_EA_) in parents were negatively correlated with ASD symptom scores on the BAPQ. In addition, multiple inherited genetic factors in parents (inhLoF, PS_EA_, PS_SZ_) were associated with parental age.

The effects of rare and polygenic variation on behavioral traits were weakly sex biased, suggesting that most genetic factors have effects on behavioral traits in both females and males. Sex-biased effects were observed for the correlation of PS_SZ_ with impairments in social communication (SCQ) and adaptive function (VABS) in sibling controls (**Fig. 5C**). Inherited risk carried by parents was associated with parental age at the birth (of probands), and the effects of rare variants (inhLOF) and PS_EA_ on parental age were paternally- and maternally-biased respectively (**Fig 5D**).

### The association of parental age with genetic risk for ASD is multifactorial

The association of multiple genetic risk factors for ASD with parental age has direct implications for understanding the mechanisms underlying parental-age effects on risk for psychiatric disorders ^25,26^. It has been established that advanced parental age is associated with ASD ^27^ and other psychiatric disorders ^28^ in offspring. We and others have demonstrated that advanced paternal age correlates with increased rates of germline mutation in offspring ^29-31^, consistent with a *de novo* mutation mechanism. An alternative model by Gratten et al. has postulated that advanced paternal age could itself be a trait that is directly influenced by inherited genetic risk that is carried by the father ^32^.

In our dataset multiple genetic risk factors for ASD were correlated with parental age (**Fig. 6**), providing evidence for both the *de novo* and inherited mechanisms. As expected, de novo SNVs were strongly correlated with paternal age in the combined dataset (r^2^paternal = 0.42, r^2^maternal = 0.27, **Supplementary Fig. 2A**), and dnLoF and dnMIS variants mirror this effect. The association of inherited risk factors with parental age was more complex, with effect sizes, directionality and gene-by-sex interactions that differ between genetic factors (**Fig. 2B**). PS_EA_ was positively correlated with parental age and maternally-biased, while inhLoF variants were negatively correlated with parental age and paternally-biased. Our results highlight how genetic mechanisms of parental-age effects may be attributable a variety of genetic factors with some having substantial effects on parental age but modest effects on ASD risk and vice versa (**Fig. 6**).

**Figure 6.**
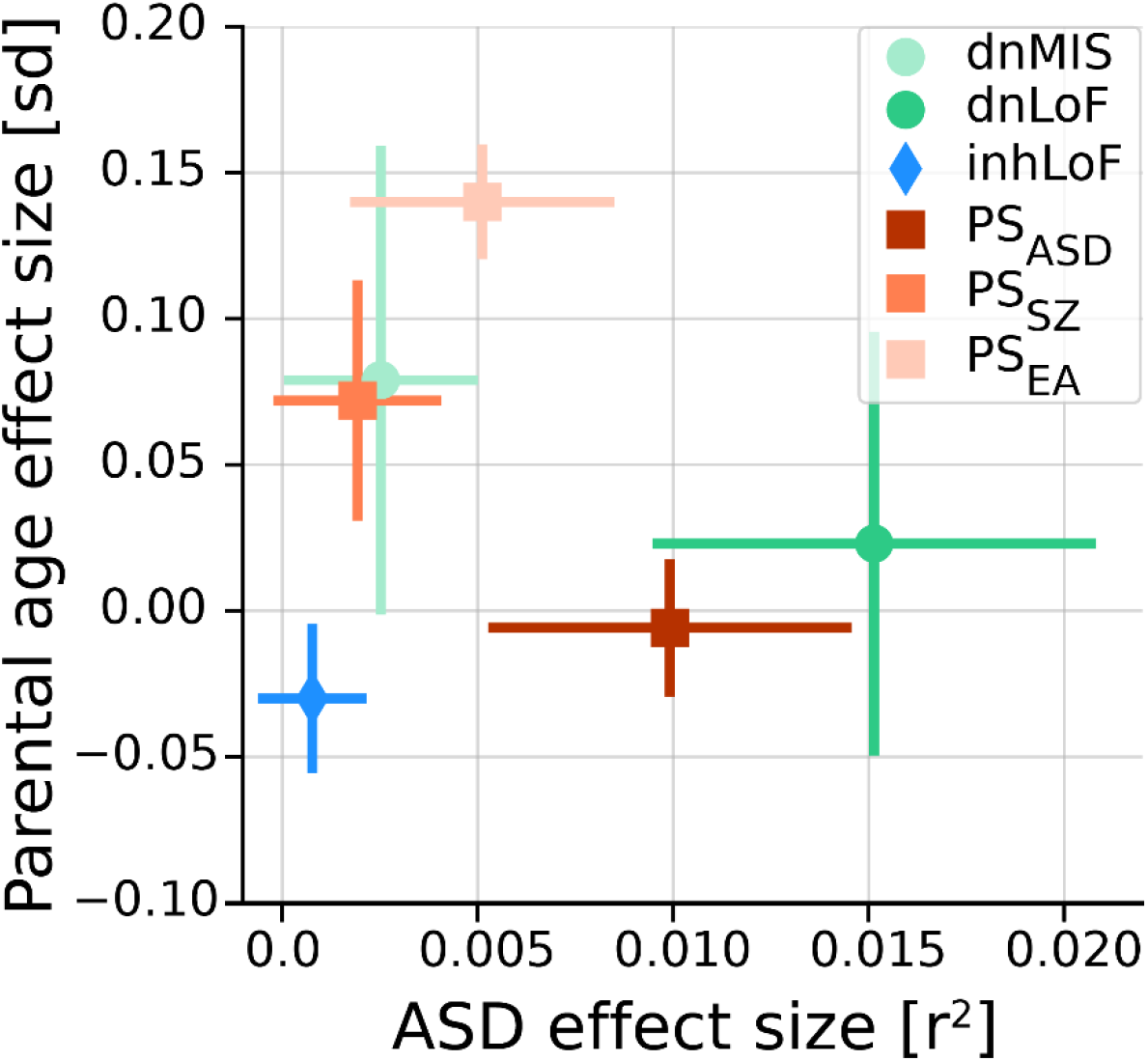
Genetic contributions to the parental-age effect on ASD risk in offspring are multifactorial. Multiple factors are correlated with both traits. Most are positively correlated with advanced parental age, and one factor (inhLoFs) is associated earlier parental age. Genetic factors with the greatest effects on ASD risk in offspring (dnLoFs, PS_ASD_) have a weak correlation with age. Conversely, factors with the strongest correlation with parental age (e.g. PSEA) have a comparatively small contribution to ASD. Effect size of EA is given as standard deviation (sd) of phenotype per unit of genetic factor. Effect size for case status is given as the Nagelkerke’s r^2^ value.

## Discussion

Whole genome analysis of a large ASD family cohort illustrates how the genetic basis of ASD consists of multiple genetic components including DNMs, rare inherited variants and polygenic scores for psychiatric and behavioral traits. Previous studies have made claims that one of several factors ^33^ has a prevailing influence. However, when the combined effects of rare and common variation are examined, we find that both have significant contributions. A composite genomic risk score accounts for additive effects of rare and common variation. However the GRS in this study is not comprehensive, and it doesn’t incorporate a variety of newly identified germline ^34,35^ and somatic ^36^ SVs. As new sequencing technologies and analytic techniques chip away at the missing heritability of ASD, new components could be incorporated into the GRS. Furthermore, as the main effects of individual genes are better understood along with non-additive effects and gene-by-sex interactions, one could further improve upon this simple model.

The genetic architectures of individual cases vary widely. Female cases have a significantly greater overall genetic load of polygenic and rare variation than male cases, demonstrating that a “female protective effect”, in which females display a higher threshold for ASD risk alleles, applies generally to all components of the genetic architecture. We find further support for differing liability thresholds in males and females as evident from a male-biased negative correlation of DNMs and polygenic scores. The negative correlation or rare-variant and polygenic risk implies that the genetic architectures in males represents a spectrum of genetic loadings that span between extremes of polygenicity and monogenic disease.

The diversity of genetic architectures that we observe contributes to phenotypic variation across the autism spectrum. We show that the multiple categories of risk influence behavioral traits in cases and in their typically-developing family members, and different genetic factors have differing patterns of trait-association. In children, DNMs were associated with deficits in motor coordination and adaptive and social behavior. Inherited risk (inhLoFs, PS_ASD_, PS_SZ_ and PS_EA_) showed differing patterns of correlation with PS_EA_ having the greatest influence on symptom severity, specifically repetitive and social behavior. Inherited risk factors for ASD carried by typically-developing parents also contributed to behavioral traits including autism symptoms (BAPQ), educational attainment and parental age, with PS_EA_ again having the greatest influence. Notably, PS_ASD_ was weakly correlated with variation in symptom severity. While surprising, there are potential explanations for this (see **supplemental material**). Perhaps the most plausible is the extreme genetic heterogeneity of SNPs that comprise PS_ASD_. The many alleles associated with case status may lack a consistent directionality with respect to specific behaviors.

Evidence for gene-by-sex interactions was detected for multiple gene-trait associations, and most interactions showed a male bias. These results provide evidence that the effects of ASD risk alleles on behavior may be stronger in males than in females, which could in part explain sex-differences in genetic load in ASD. However, it may be premature to conclude that the effects of ASD risk alleles in general are muted in females given that two major factors (DNMs and PSASD) did not show strong gene-by-sex effects on behavioral traits. Also, there has been suggestive evidence from exome sequencing that sex biases act in both directions with some genes having mutations more frequently in females while others appear to have a male bias ^37^. Also, it is likely that social factors or biases in clinical ascertainment may contribute to a male preponderance of ASD ^38^. Further investigation of sex bias in the effects of genetic factors on behavior in the general population could help to elucidate what traits in females are most strongly influenced by genetic risk for ASD and could help to determine whether the “silent” (undiagnosed) subset of females with high genetic load may exhibit different clinical features and potentially receive different diagnoses.

Lastly, our results provide new insights into the genetic mechanisms of parental-age effects on ASD risk in offspring ^39^. One mechanism is *de novo* mutation, a major component of ASD risk for which paternal age has a direct influence ^29,40^. Another is the mechanism of inherited risk in which parental age is not a direct cause of ASD in offspring, but is instead a behavioral trait in adults that is influenced by the risk alleles they carry ^32^. This study provides genetic evidence for the inherited mechanism, and further highlights the sex-biased effects of genes on behavior. For example, a genetic predictor of high educational attainment (PS_EA_) is associated with later parenthood, particularly for women. We confirmed in our dataset that education and parental age ^41^ were more strongly correlated for mothers (r^2^ = 0.06, p = 3.5×10^−52^) than for fathers (r^2^ = 0.03 p=1.3×10^−23^). Thus, inherited mechanisms of maternal-age effects may involve a dependency between the level of education a woman seeks and the age at which she has children. Conversely, a genetic predictor of low educational attainment (inhLoF) was negatively correlated with parental age, specifically for men. This result is consistent with an increased susceptibility of males to rare-variant risk. Thus rare variants of large effect in fathers could explain an increased risk for ASD in the children of parents who are very young ^42^. Likewise, previous reports of a U-shaped effect of father’s age on risk for developmental impairments in offspring ^43^ may be attributable to the combined effects of inhLoF, DNMs and PS_EA_.

The results described here highlight how an integrated analysis of multiple genetic factors can improve our understanding of the genetic basis of ASD. While most of the heritability of ASD remains unexplained, the expanding arsenal of sequencing platforms and methods of variant detection promise to expand the range of genetic factors that can be captured from a genome. The growing cohorts of ASD ^19^ as well as individual rare diseases ^44^ promise to improve knowledge of the effects of risk alleles on psychiatric traits and how their combined effects determine clinical outcome.

## Data Availability

WGS data from the SSC and Exome and SNP genotyping data from SPARK are available from the Simons Foundation Autism Research Initiative (SFARI) Summary genetic data including individual counts for dnLoF, dnMis, inhLoF and polygenic scores for all subjects in this study and input data files for all analysis code are also available from SFARI. WGS data from the REACH project are available from the NIMH Data Archive (NDA) including the structural variant callset, and raw sequence (FASTQ), alignment (BAM) and variant call (VCF) files from the REACH cohort.
Analysis code for the major statistical genetic analyses in the paper is available as a Google Colab notebook on Github https://github.com/sebatlab/Antaki2021. Analysis code is provided for the following.
Variance Explained by Genetic Risk Factors (Fig 2A)
Negative correlations of rare and common genetic risk (Fig. 3)
Gene/phenotype correlations (Fig. 5) 

https://www.sfari.org/resource/autism-cohorts

https://nda.nih.gov/edit_collection.html?id=2019

## Acknowledgements

We wish to thank the families who participated in genetic studies of the REACH, SSC and SPARK cohorts. We would also like to thank W. Pfeiffer, M. Gujral, the San Diego Supercomputer Center, and Amazon Web Services for hosting the computing infrastructure necessary for completing this project. We wish to thank Wendy Chung and SFARI for providing data and materials for validation of DNM calls. This work was supported by grants to J.S. from the Simons Foundation Autism Research initiative (SFARI 606768), National Institutes of Health (MH113715, MH119746, 1MH109501) and the Escher fund for Autism (20171603) and grant to C.N. from the National Institutes of Health (MH106595). D.A. was supported by A T32 training grant from the NIH (GM008666). M.K. was supported by a Rubicon grant from the Netherlands Organization for Scientific Research (NOW 45219212).

## Data and Code availability

WGS data from the SSC and Exome and SNP genotyping data from SPARK are available from the Simons Foundation Autism Research Initiative (SFARI) (https://www.sfari.org/resource/autism-cohorts). Summary genetic data including individual counts for dnLoF, dnMis, inhLoF and polygenic scores for all subjects in this study and input data files for all analysis code are also available from SFARI. WGS data from the REACH project are available from the NIMH Data Archive (NDA) including the structural variant callset, and raw sequence (FASTQ), alignment (BAM) and variant call (VCF) files from the REACH cohort (https://nda.nih.gov/edit_collection.html?id=2019).

Analysis code for the major statistical genetic analyses in the paper is available as a Google Colab notebook on Github https://github.com/sebatlab/Antaki2021. Analysis code is provided for the following.

*Variance Explained by Genetic Risk Factors (****Fig 2A***)

*Negative correlations of rare and common genetic risk (****Fig. 3***)

*Gene – phenotype correlations (****Fig. 5***)

## Online Methods

### Datasets

The sample was comprised of three datasets, including whole genome sequencing of cohorts from the REACH project at UCSD (https://sebatlab.org/reach-project) and the Simons Simplex Collection (SSC) and a dataset of exomes and SNP genotyping from the SPARK study ^19^ (Supplementary Table 1). The combined sample of 11,313 ASD families consisted of a total 37,375 subjects including 12,270 cases, 5,190 TD siblings and 19,917 parents (Supplementary Table 1). See data and code availability for details on data access.

### Processing of DNA sequence data

Each of the three datasets consisted of Illumina paired-end sequence data which were processed by BWA alignment and variant calling using GATK best practices. Specific differences between datasets include library prep (PCR vs PCR free, WGS vs exome) and differences in software version. Details are provided in the sections below. Analysis was carried out with SNP, indel and SV variant calls mapped to GRCh38. All calls were generated from sequence aligned to GRCh38 or lifted over from GRCh37 to GRCh38 prior to annotation and analysis.

### REACH cohort

Whole genome sequencing was performed on blood-derived genomic DNA as described in our previous publication ^45^. Standard quality control steps were carried out to to ensure proper relatedness and genetic sex concordance with the sample manifest. Sequencing reads were aligned to the GRCh37 reference genome using bwa-mem (v0.7.12). Subsequent processing of the alignments followed GATK Best Practices guidelines including sorting, marking duplicate reads, indel realignment, and base quality score recalibration.

To ensure functional equivalency with other cohorts in our dataset, we applied the same SNV/indel variant calling pipeline used on the SSC cohort (see SSC section below for details). We utilized GATK HaplotypeCaller (v4.1) to first call SNVs and indels in individual samples. GRCh38 GVCFs were then combined using CombineGVCF and jointly genotyped. Variants were then subjected to variant quality score recalibration (VQSR). The VQSR model was trained with the parameter “maxGaussians=8” for SNVs and “maxGaussians=4” for indels. Variant scores were recalibrated with the truth sensitivity level of 99.8% for SNVs and 99.0% for indels. Sample-level filtering converted genotypes to noncalls (“./.”) if the GQ < 20 or the DP < 10. Lastly, before proceeding with variant annotation, variants were lifted over from GRCh37 to GRCh38 with the GATK LiftoverVcf command.

### Simons Simplex Cohort

Whole genome sequencing was performed at the New York Genome Center (NYGC) on an Illumina HiSeq X10 sequencer using 150bp paired-end reads to an average depth of 40x. Reads were aligned to the GRCh38 reference genome using bwa-mem with subsequent processing of alignments in line with GATK Best Practices for functional equivalence.

SNV and indel joint genotypes were provided by the NYGC (dated 2019-03-21). SNV and indel calling was performed using GATK (v3.5). Variant discovery implemented HaplotypeCaller in GVCF mode. Subsequent GVCFs were combined in batches of approximately 130 samples (average) using CombineGVCFs. The remaining combined GVCFs were jointly called creating a genotype matrix of 9209 samples. The jointly genotyped VCF was then subjected to variant score recalibration using a Variant Quality Score Recalibration (VQSR) model. The VQSR model was trained with the parameter “maxGaussians=8” for SNVs and “maxGaussians=4” for indels. Variant scores were recalibrated with the truth sensitivity level of 99.8% for SNVs and 99.0% for indels. Sample-level flagging of low quality, defined as GQ < 20 or DP < 10. Analysis was carried out on the “masked” VCFs where genotypes that failed to meet the above sample-level filtering were converted to missing calls (“./.”). Only variants that had “PASS” entries in the FILTER column were considered for inherited analysis. Further details on the generation of the SSC SNV and indel joint calls can be found in the PDF accompanying the data release from the Simons Foundation.

### SPARK Cohort

The publically available SPARK dataset consists SNP genotyping (Illumina global screening array GSA-24v1-0) and exomes (IDT xGen capture sequenced on the Illumina NovaSeq 6000 using 2/S4 flow cells). Imputation of SNP genotypes was performed using the RICOPILI pipeline (https://sites.google.com/a/broadinstitute.org/ricopili/imputation) ^46^.

Downstream processing of exome data was performed as follows. Per-sample GVCFs were obtained from the SPARK September 2019 data release (an update to the original release in which duplicate reads were marked). GVCFs had been generated with GATK v4.1.2.0 HaplotypeCaller from CRAM files aligned to GRCh38 with bwa-mem. Joint genotyping of SNP and indel variant calls was performed at UCSD in batches of 100 families the same GATK pipeline that was used for the REACH and SSC WGS. Briefly, variant scores were recalibrated with the truth sensitivity level of 99.8% for SNVs and 99.0% for indels. Sample-level flagging of low quality, defined as GQ < 20 or DP < 10. Analysis was carried out on the “masked” VCFs where genotypes that failed to meet the above sample-level filtering were converted to missing calls (“./.”). Variants with “PASS” in the FILTER column were retained for analysis. Likewise, indel calls with QD < 7.5 were omitted.

### Principal Components (PCs) Calculation

Genotype data was LD pruned to a set of 100,370 unambiguous markers with minor allele frequency > 5% in PLINK 1.9, using the ╌indep-pairwise command with a 200 variant window, shifting the window 100 variants at a time, and pruning variants with r^2^ > 0.2. KING version 2.2.4 (https://doi.org/10.1093/bioinformatics/btq559) was used to identify a set of unrelated individuals (1^st^ and 2^nd^ degree related individuals removed). PCs were calculated in the unrelated individuals based on LD pruned data using FlashPCA2 (https://doi.org/10.1093/bioinformatics/btx299) and related individuals were then projected onto the PCs.

### Polygenic Risk Score (PRS) calculation

Polygenic scores (PS_ASD_) and (PS_SZ_) were calculated on based on current ASD and SZ summary statistics from the psychiatric genomics consortium (https://www.med.unc.edu/pgc/download-results/). In the case of ASD, PGC summary statistics were recalculated by J.G. from weighted meta-analysis of all study-wise summary statistics in Grove et al. ^18^ excluding the SSC dataset used in this study. PS_EA_ was calculated from summary statistics of the recent GWAS meta-analysis of educational attainment by Lee et al ^47^. Polygenic scores were calculated using PRSice version 2.3.0 (https://doi.org/10.1093/gigascience/giz082). Only unambiguous variants with MAF > 1% in the reference dataset were included. Variants were LD clumped over a 250kb window with an r^2^ value of 0.1. PS were calculated at multiple p-value thresholds (0.01-0.9) to determine the optimal threshold. The best fitting PS for each trait was selected based on significance level of a TDT test carried out in autism cases (p-value threshold = 0.1 for ASD and SCZ, p-value threshold = 0.05 for EA). The best fitting PS was carried forward for all subsequent statistical analyses.

### SV calling

SV calls were only produced for the whole genome datasets: REACH and SSC. Our SV calling and filtering workflow has been described in detail in our previous publication (Brandler et al. 2018). Briefly, we ran ForestSV, LUMPY, and Manta on each sample calling deletions and duplications. ForestSV mainly relies on coverage as a feature to call SVs, resulting in segmented calls for large events that span repetitive elements such as segmental duplications. Because of this, we applied a stitching algorithm to ForestSV calls, combining calls of the same SV class if they were ≤10kb apart. As a preliminary filter, we omitted any variant that overlapped more than ⅔ of the SV length to centromeres, telomeres, segmental duplications, regions with low mappability with 100bp reads, antibody parts, T-cell receptors, and other assembly gaps.

The resulting calls were genotyped using SV2 and SVTyper within each sample. SVs and genotypes were then collapsed for overlapping calls. The collapsing algorithm first prioritized the breakpoint confidence intervals if both the start and end confidence intervals provided by LUMPY and/or Manta overlapped. For ForestSV calls, the confidence interval was defined as +/-100bp from the start and end positions. The consensus position determined for a collapsable cluster was determined by the SV position with the highest number of overlaps. In the case of a tie, the median position was recorded. This method allows for collapsing of common SV that “tile” across a region, which rarely occurs outside of variable regions such as the HLA locus. The resulting calls were then subject to a further round of collapsing, this time reducing calls to a consensus position if they overlapped 80% reciprocally with each other. This method was applied recursively until no more calls could be collapsed. As for confidence interval collapsing, the consensus position reported was the SV with the highest number of overlaps. Variant level genotype likelihood scores were generated with SV2 by pooling all features from REACH and SSC samples. If the SV2 variant score was not “PASS” then the SVTyper or Manta genotypes were recorded, as previously described (Brandler et al. 2018). Samples without a genotype call were considered as missing (“./.”).

### DNM calling

DNMs (DNM) were called using the synthDNM software ^48^. SynthDNM is a random forest (RF)-based classifier which uses only a pedigree file (PED/FAM) and VCF files as input and can be readily optimized for different technologies or variant calling pipelines. For WGS datasets (REACH and SSC) we used the default SynthDNM classifier (SSC1 GATK) which was trained on GATK variant calls from >30X Illumina WGS data. This default classifier had high accuracy (AUC = 0.997) for detecting a truth-set of orthogonally validated de novo SNVs and indels from SSC ^48^. For the exome dataset (SPARK) we trained an additional four classifiers, one for each set of variant calls: DeepVariant, WeCall, SPARK GATK, and SSC GATK. To maximize sensitivity while controlling for false positives, we retained DNMs if they were called by three out of the five classifiers. To further confirm the accuracy of SPARK DNM calls, we compared the de novo SNV and indel calls on the SPARK dataset to a set of validated DNMs that were confirmed by Sanger sequencing from a previous pilot study (Feliciano et al., 2019). For SNVs, the recall rate for SNVs ranged from 92.6% to 98.2% (N=117), while for indels the recall range from 98.6 to 100% (N=107). For further details of the methodology and performance of SynthDNM, refer to our companion paper ^48^.

De novo SVs were defined as events with heterozygous genotypes in offspring and homozygous reference calls in parents. We only considered variants that passed the stringent “DENOVO_FILTER” filter produced by SV2 ^49^. We applied our standard filtering guidelines detailed below to omit variants present in regions known to produce spurious calls. We also supplemented our de novo calls with the de novo CNV calls generated from microarrays in SSC samples from Sanders et al. Neuron 2015 since many of these calls are likely to be missed by paired-end SV callers. We then manually inspected the list of de novo SVs and stitched calls together if they were separated by segmental duplications greater than 10kb (the maximum stitching requirement for ForestSV calls detailed in the section below).

### Variant Annotation

Variant Effect Predictor (VEP) v97 along with transcript annotations from Gencode v31 were used in annotation of SNVs and indels. Variants were flagged as “LoF’’ if the functional consequence one of the following: “transcript_ablation”, “splice_acceptor_variant”, “splice_donor_variant”, “stop_gained”, “frameshift_variant”, “stop_loss”, “start_loss”. LoF variants exclusive to nonsense mediated decay transcripts were omitted from subsequent analysis. SVs were annotated by overlap to exons and proximal cis-regulatorly elements including 5’UTRs, transcription start sites, and fetal brain promoters. Since the list of annotated proximal cis-regulatory elements were in GRCh37, we lifted over the GRCh38 SV calls to GRCh37 for all subsequent analysis.

We attributed gnomAD LOEUF scores (v2.1.1) to each LoF variant and CRE-SV. If a variant overlapped more than one gene, as in the case for large SVs, we recorded the minimum (most constrained) LOEUF score to that variant. Constraint was quantified for missense variants using the “Missense Badness, PolyPhen-2, and Constraint” (MPC) scores (bioRxiv https://doi.org/10.1101/148353). These scores are available for the GRCh37 build of the human genome and were transposed to GRCh38 for analysis. Missense variants without MPC scores due to updates to the reference genome were not used in subsequent analysis. The recommended cutoffs to enrich for the top tier of constraint (LOEUF < 0.37; MPC > 2) was applied to de novo and rare inherited LoF variants.

### Association tests

#### Defining the variant types to be tested

Unless otherwise specified, all target categories consisted of private variants in which the alt allele was present in only one family in this study, and target categories included only variants in functionally constrained genes as defined below.

**MIS** variants were defined as all private missense SNVs with MPC scores > 2. **LoF** variants were defined as variant calls that were predict to result in loss of protein function (truncation of a protein) and included stop-gain, frameshift, splice site and exonic deletion in a loss-of-function intolerant gene defined as LOEUF < 0.37, per recommendations from gnomAD. SVs that intersected more than one gene were assigned minimum LOEUF score (corresponding to the most constrained gene).

Additional filters were applied to the rare inherited variants (inhLoF). We included only variants with a “PASS” entry in the “FILTER” column and we removed SNVs and indels with ≥ 5% missing calls across the cohorts and any variant that was observed to have an allele frequency more than 1% in gnomAD (v2.1.1). For private variants in the SPARK WES dataset, we applied one additional filter removing variants with QD scores less than 7.5. For SVs, we included only exonic SVs >50bp in length and CRE-SVs ≥ 2.5kb that passed the “DENOVO_FILTER” from the SV2 software which is a stringent filter recommended for ultra-rare variants.

#### De Novo Association

The burden of damaging DNMs was compared between cases and controls by a two sample independent T-test reporting the two-sided p-values. For results and the full list of damaging DNMs please refer to **Supplementary Table 2**.

#### Inherited Rare Association

The number of transmissions and nontransmissions from parent to offspring was obtained using plink’s “╌tdt poo” command (v1.9). Pooling of transmission and nontransmission counts for the transmission disequilibrium test (TDT) was done using the pytdt python package (https://github.com/sebatlab/pytdt). This package takes as input a data table containing a unique variant ID and counts for transmissions and nontransmissions in fathers and mothers for both cases and controls. Pytdt performs the pooling or group-wise analysis of private LoF variants and CRE-SVs by summing the counts of transmissions and non-transmissions for all variants encompassing a group. We also conditioned the TDT according to damaging DNM burden in the offspring. The binomial test is used for statistical significance of transmission distortion of private variants to cases or controls separately. The package also reports odds ratios, confidence intervals, and other statistics commonly used for TDT analysis. For a summary of the TDT results and a list of all the private variants tested in the analysis refer to **supplementary table 3**.

#### Polygenic TDT

Per methods from Weiner et al. ^17^ trio-based association of polygenic scores (PS_ASD_, PS_SZ_, PS_EA_) with ASD was tested with the polygenic TDT (pTDT), which tests the significance of the deviation of the child PS from the average PS of the parents.

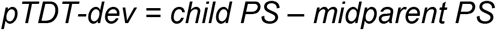

P-value was then calculated with a one sample T-test of pTDT-dev (**Fig. 1C**) with a population mean of 0. All polygenic risk scores and derivatives of them are reported in the sample manifest (**Supplementary Table 1**) and results of the pTDT are reported in **Supplementary Table 4**.

#### Calculating the composite risks scores of multiple factors

We used multivariable regression to capture the combined effects of multiple genetic factors on case status. For rare variant factors, the predictor variables in the model consisted of rare variant burden counts for dnMIS, dnLoF and inhLoF. For polygenic scores PS_ASD_, PS_SZ_ and PS_EA_, the predictor variables consisted of the pTDT-dev values of the trios. To calculate a composite genetic risk score, each predictor variable was first residualized for PCs and sex. Then estimates were calculated from a generalized linear model as follows

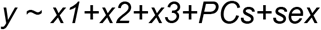

where y is case status and x1, x2 and x3 are residualized predictor variable for three genetic factors. PCs for all regression models consisted of the first 10 principal components from the PCA. Then the composite risk score (RS) is calculated using *r* as

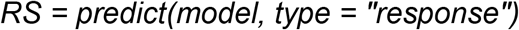

Each RS was than standardized by Z-transformation. Predictor variables (x1, x2, x3, etc.) for each risk score consisted of

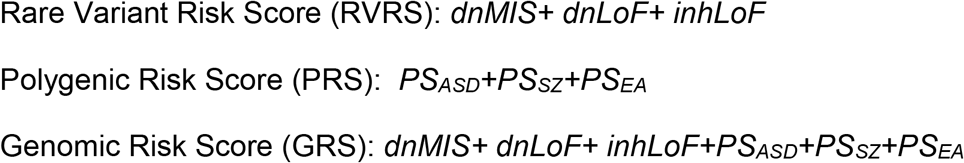

To compare the effect sizes on case status for the genetic factors and the composite risk scores (**Fig. 2B**), Nagelkerke’s r^2^ values were calculated for each of the residualized predictor variables and for each composite risk score.

#### Pairwise correlations of rare variants and polygenic risk

To test the correlations between rare variants and polygenic risk, we constructed pairwise linear models

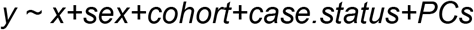

Where the variable y is a polygenic score (PS_ASD_, PS_SZ_, PS_EA_ or PRS) and x is a measure of rare variant load (dnLoF, dnMIS, inhLoF or RVRS). Gene-by-sex interaction was then tested in the following model.

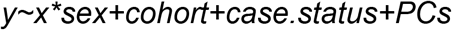

**Supplementary Table 5** contains the full results for all pairwise correlation of rare and polygenic risk conditioned on sex.

### The effects of genetic factors on behavioral traits

The effects of genetic factors on behavioral traits was investigated in the SSC and SPARK cohorts using clinical phenotype data available from SFARI (see data and code availability). To eliminate confounders due to ancestry only subjects of european ancestry confirmed by PCA were included. Clinical measures of ASD symptoms and related behaviors were selected that were available for cases, typically-developing sibling or parents. Phenotype measures consisted of the summary scores from the developmental coordination disorder questionnaire (DCDQ) of motor function and the Repetitive Behavior Scale (RBS) that were available on cases; the Vineland Adaptive Behavior Scale (VABS), Social Communication Questionnaire (SCQ) and Social Responsiveness Scales (SRS) that were available on both cases and TD siblings. Behavioral phenotypes available on parents included the Broad Autism Phenotype Questionnaire (BAPQ), parental educational attainment (from the background history questionnaire) and parental age at birth (for the children with ASD diagnosis). Phenotype measures that were available for both the SSC and SPARK cohorts were normalized within cohort by Z-transformation, then combined, and cohort was included as a covariate in the downstream analyses. A summary of the samples available for each phenotype measures for each subject are provided in the master sample manifest (Supplementary Table 2).

Association of genetic factors with developmental traits was tested by linear regression controlling for sex, cohort and principal components. In addition a gene-by-sex interaction was tested to determine if genetic effects on cognitive traits differed for males and females. Phenotypes in offspring (cases and siblings) were tested using the model

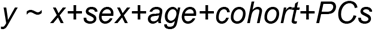

Where y is the phenotype variable and x is the genetic factor (DNMs, IRVs and PSes). In addition a gene-by-sex interaction was then tested in this model.

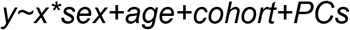

## Supplementary Materials and Results

### Contents

#### 3. Supplementary Tables

Supplementary Table 1: Sample Manifest with individual-level variant counts and polygenic scores

Supplementary Table 2: DNM Analysis (Results from Fig. 1A) and lists of DNMs detected in this study

Supplementary Table 3: Analysis of inherited rare variants (Results from Fig. 1B) and lists of inherited rare variants detected in this study (inhLoF, LoF-SV and CRE-SV)

Supplementary Table 5: Polygenic TDT (Results from Fig 1.C)

Supplementary Table 6: Genetic Correlations of Rare Variants and Polygenic Scores Results for Figure 4B)

Supplementary Table 7: Correlation of Genetic Factors with Behavioral Traits in Cases, Sibling Controls and Parents (Results from Fig. 5)

#### 1. Supplementary Results

##### 1.1 The influence of sex on the transmission of inherited rare variants in families

Linear regression analysis of trios controlling for cohort, sex and case status shows a negative correlation of DNMs and inhLoF (**Supplementary Figure 1A**, P = 0.03) and no significant gene by sex interaction. This result is consistent with a liability threshold model, in this case with effects that are similar in males and females. These results are consistent with familial and sporadic ASD having distinct etiologies (i.e. cases that carry damaging DNMs have less inhLoF risk and vise versa).

Based on the model of a female protective effect, we have proposed previously that the differential susceptibility of males and females could result in a bias in the transmission of genetic risk in ASD families ^8^. Specifically, we hypothesized that inherited risk might show a biased transmission from the “protected” parent (mothers) to “susceptible” offspring (male cases). However, in this study we did not observe evidence that inhLoF variants were maternally biased, and we did not observe evidence of the predicted mother to son bias (**Supplementary Figure 1B**) in the transmission of inhLoF variants. While we did observe a trend of increased mother-to-daughter transmission, consistent with there being a maternal bias of inherited risk in female cases, the difference was not statistically significant. Based on analysis of gene-phenotype correlations we do find some evidence for a paternal bias in the effects of inhLoF variants on parental behavior (Fig. 5C). However, sex did not have a strong effect on the transmission of inhLoF variants in families. Our results suggest that both fathers and mothers contribute substantially to inhLoF risk.

##### 1.2 Apparent discrepancies in the associations of polygenic scores with case status and symptom severity

###### PS_ASD_

PS_ASD_ is one of the major genetic predictors of case status in this study (**Fig. 2A**), but it does not correlate strongly with the measures of symptom severity (**Fig. 5**). PS_ASD_ shows a nominally-significant correlation with social communication (SCQ) in cases but does not show statistical correlation with any other measures including repetitive behavior (RBS) and social responsiveness (SRS). PS_ASD_ was also a comparatively weak correlate of behavioral traits in the UK Biobank compared with polygenic scores for other psychiatric disorders ^50^. There are a few potential explanations for the weak effects of PS_ASD_ on symptom severity. **(1)** One explanation is the high genetic heterogeneity (and the corresponding phenotypic heterogeneity) within the autism spectrum. Studies of CNVs have shown that reciprocal deletions and duplications of the same genes, for example CNVs at 16p11.2 ^51-53^ and 1q21.1 ^54^, both associate with ASD, but the deletion and duplication alleles have very different effects on quantitative traits. We hypothesize that the effects of the many common ASD risk alleles may have similarly heterogeneous effects, such that all are associated with case status but have inconsistent directionality of effects on specific quantitative measures of behavior. **(2)** Another explanation may be limited statistical power of existing ASD GWAS. Sample sizes for ASD are smaller than for other disorders such as schizophrenia and bipolar disorder, and thus summary statistics could be less informative, but given that PS_ASD_ is the polygenic score of strongest effect, the latter hypothesis is less satisfying than the former.

###### PS_EA_

Interestingly PS_EA_ shows opposing effects: a positive correlation with case status (**Fig 1C**) and negative correlations with symptom severity (**Fig. 5**). Thus, a genetic predictor of high educational attainment predisposes to ASD, a fact that has led to much discussion regarding relationship of high intelligence to clinical features of autism ^55^. However that same genetic predictor had protective effect on social (SCQ, SRS) and repetitive behavior (RBS). Thus, the relationship of PS_EA_ to ASD risk is complex. A few potential mechanisms could explain the observed patterns. PS_EA_ may contribute to a clinically-distinct subtype of high-functioning ASD, and PS_EA_ could be positively correlated with a subtype of social deficits that were not tested in this study. Indeed Grove et al has reported that the effect size for PS_EA_ was strongest in the “Asperger syndrome” clinical subtype ^18^. **(2)** Notably the single strongest phenotypic correlate of PS_EA_ was in fact a risk factor for ASD: parental age. It is conceivable that increased PS_EA_ in parents could result in increased age and increased rates of DNMs. However the pTDT is controlled for such parental age effects. An over-transmission of PS_EA_ to cases is consistent with the former hypothesis.

###### Supplementary Figures

**Supplementary Figure 1.**
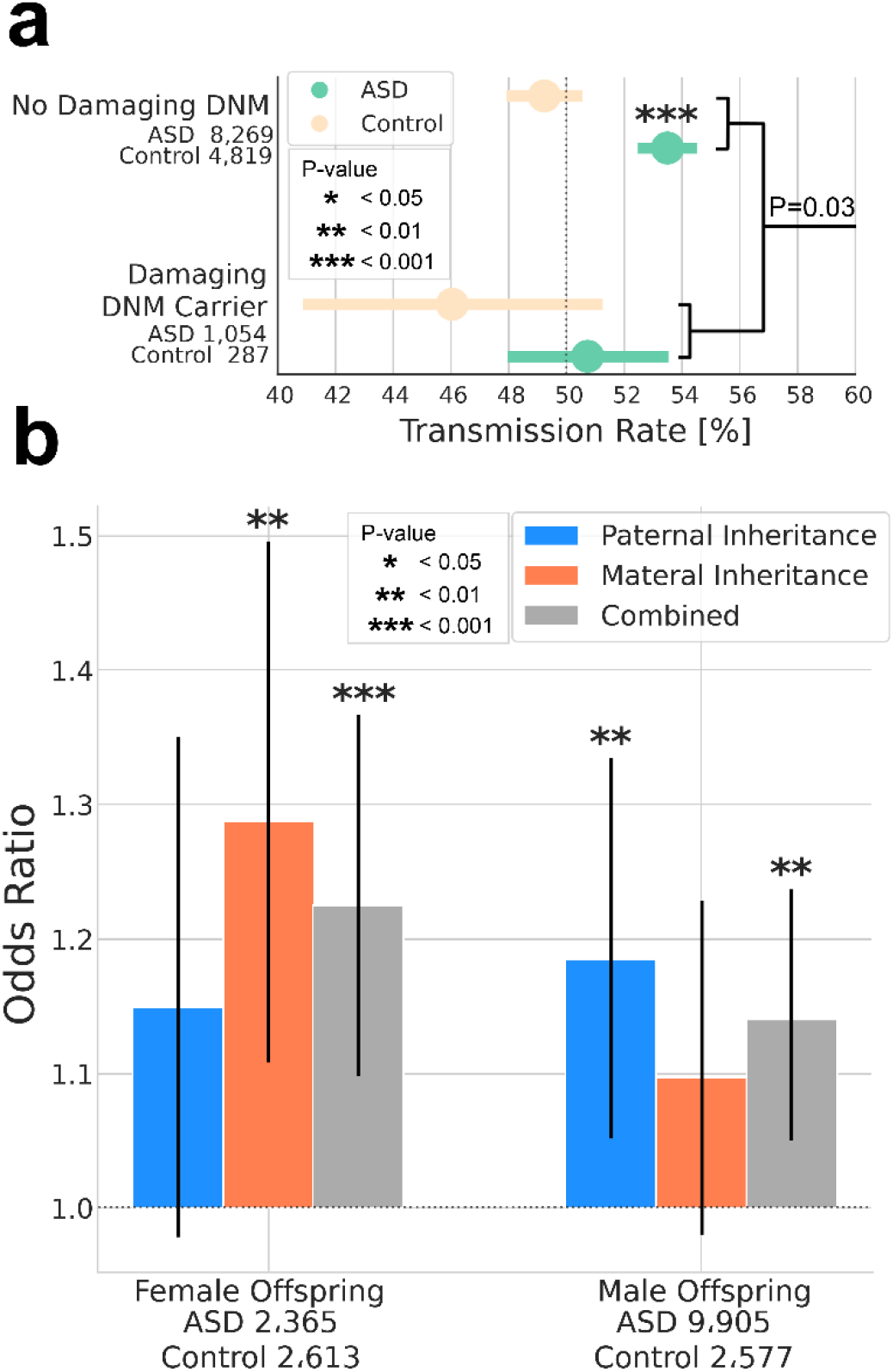
The combined effects of dnLoF, inhLoF and sex on the transmission of rare variants in families. (a) A significant liability threshold for rare variants was evident based on a negative correlation of dnLoF and inhLoF (linear regression P = 0.03), and this effect did not differ significantly by sex. (b) Case-control odds ratios were compared for the transmission rates in families by sex (father-daughter, father-son, mother-daughter, mother-son). Both maternal and paternal rare variants contribute to ASD with a significant over-transmission from mother to daughter and from father to son. We did not observe a significant sex bias in the transmission of rare variants in families. In particular, we did not observe an enriched transmission from mother to male cases as we have previously hypothesized ^8^.

**Supplementary Figure 2.**
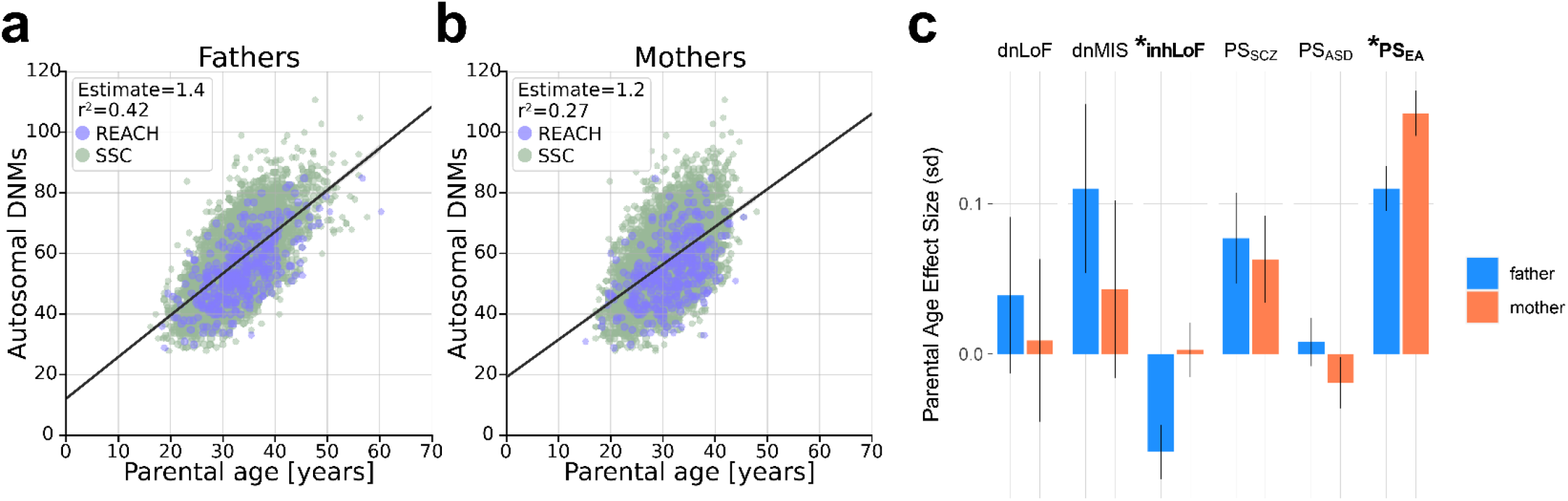
Correlation of *de novo* and inherited risk with parental age. Correlation of total autosomal *de novo* SNVs with age of (**a**) fathers and (**b**) mothers. (**c**) Correlations of each genetic factors with paternal and maternal age, “*” denotes significant gene-by-sex interactions (P<0.05), see also figure 5D.

## Notes

### Competing Interest Statement

The authors have declared no competing interest.

### Author Declarations

This study was carried out as part of protocol 101413 approved by the Institutional Review Board of the UCSD Human Research Protections Program

## References

1. Sebat, J. et al. Strong association of de novo copy number mutations with autism. Science 316, 445–9 (2007).

2. Iossifov, I. et al. The contribution of de novo coding mutations to autism spectrum disorder. Nature 515, 216–21 (2014).

3. Grove, J. et al. Common risk variants identified in autism spectrum disorder. bioRxiv (2017).

4. Sebat, J., Levy, D.L. & McCarthy, S.E. Rare structural variants in schizophrenia: one disorder, multiple mutations; one mutation, multiple disorders. Trends Genet 25, 528–35 (2009).

5. Bergen, S.E. et al. Joint Contributions of Rare Copy Number Variants and Common SNPs to Risk for Schizophrenia. Am J Psychiatry, appiajp201817040467 (2018).

6. Davies, R.W. et al. Using common genetic variation to examine phenotypic expression and risk prediction in 22q11.2 deletion syndrome. Nat Med 26, 1912–1918 (2020).

7. Lim, E.T. et al. Rare complete knockouts in humans: population distribution and significant role in autism spectrum disorders. Neuron 77, 235–42 (2013).

8. Zhao, X. et al. A unified genetic theory for sporadic and inherited autism. Proc Natl Acad Sci U S A 104, 12831–6 (2007).

9. Werling, D.M. & Geschwind, D.H. Recurrence rates provide evidence for sex-differential, familial genetic liability for autism spectrum disorders in multiplex families and twins. Mol Autism 6, 27 (2015).

10. Robinson, E.B., Lichtenstein, P., Anckarsater, H., Happe, F. & Ronald, A. Examining and interpreting the female protective effect against autistic behavior. Proc Natl Acad Sci U S A 110, 5258–62 (2013).

11. Sanders, S.J. et al. Insights into Autism Spectrum Disorder Genomic Architecture and Biology from 71 Risk Loci. Neuron 87, 1215–33 (2015).

12. Desachy, G. et al. Increased female autosomal burden of rare copy number variants in human populations and in autism families. Mol Psychiatry 20, 170–5 (2015).

13. Jacquemont, S. et al. A higher mutational burden in females supports a “female protective model” in neurodevelopmental disorders. American Journal of Human Genetics 94, 415–25 (2014).

14. De Rubeis, S. et al. Synaptic, transcriptional and chromatin genes disrupted in autism. Nature 515, 209–15 (2014).

15. Brandler, W.M. et al. Paternally inherited noncoding structural variants contribute to autism. bioRxiv (2017).

16. Krumm, N. et al. Excess of rare, inherited truncating mutations in autism. Nat Genet 47, 582–8 (2015).

17. Weiner, D.J. et al. Polygenic transmission disequilibrium confirms that common and rare variation act additively to create risk for autism spectrum disorders. Nat Genet 49, 978–985 (2017).

18. Grove, J. et al. Identification of common genetic risk variants for autism spectrum disorder. Nat Genet 51, 431–444 (2019).

19. Spark Consortium. SPARK: A US Cohort of 50,000 Families to Accelerate Autism Research. Neuron 97, 488–493 (2018).

20. Werling, D.M. The role of sex-differential biology in risk for autism spectrum disorder. Biol Sex Differ 7, 58 (2016).

21. Falconer, D.S. Inheritance of Liability to Certain Diseases Estimated from Incidence among Relatives. Annals of Human Genetics 29, 51-& (1965).

22. Reich, T., Morris, C.A. & James, J.W. Use of Multiple Thresholds in Determining Mode of Transmission of Semi-Continuous Traits. Annals of Human Genetics 36, 163-& (1972).

23. Robinson, E.B. et al. Genetic risk for autism spectrum disorders and neuropsychiatric variation in the general population. Nat Genet 48, 552–5 (2016).

24. Satterstrom, F.K. et al. Large-Scale Exome Sequencing Study Implicates Both Developmental and Functional Changes in the Neurobiology of Autism. Cell 180, 568–584 e23 (2020).

25. Reichenberg, A. et al. Advancing paternal age and autism. Arch Gen Psychiatry 63, 1026-32 (2006).

26. Malaspina, D. et al. Advancing paternal age and the risk of schizophrenia. Arch Gen Psychiatry 58, 361–7 (2001).

27. Croen, L.A., Najjar, D.V., Fireman, B. & Grether, J.K. Maternal and paternal age and risk of autism spectrum disorders. Arch Pediatr Adolesc Med 161, 334–40 (2007).

28. Brown, A.S. et al. Paternal age and risk of schizophrenia in adult offspring. Am J Psychiatry 159, 1528–33 (2002).

29. Michaelson, J.J. et al. Whole-genome sequencing in autism identifies hot spots for de novo germline mutation. Cell 151, 1431–42 (2012).

30. Kong, A. et al. Rate of de novo mutations and the importance of father’s age to disease risk. Nature 488, 471–5 (2012).

31. Goriely, A. & Wilkie, A.O. Missing heritability: paternal age effect mutations and selfish spermatogonia. Nat Rev Genet 11, 589 (2010).

32. Gratten, J. et al. Risk of psychiatric illness from advanced paternal age is not predominantly from de novo mutations. Nat Genet 48, 718–24 (2016).

33. Gaugler, T. et al. Most genetic risk for autism resides with common variation. Nat Genet 46, 881–5 (2014).

34. Trost, B. et al. Genome-wide detection of tandem DNA repeats that are expanded in autism. Nature 586, 80–86 (2020).

35. Mitra, I. et al. Patterns of de novo tandem repeat mutations and their role in autism. Nature 589, 246–250 (2021).

36. Sherman, M.A. et al. Large mosaic copy number variations confer autism risk. Nat Neurosci 24, 197–203 (2021).

37. Turner, T.N. et al. Sex-Based Analysis of De Novo Variants in Neurodevelopmental Disorders. Am J Hum Genet 105, 1274–1285 (2019).

38. Russell, G., Steer, C. & Golding, J. Social and demographic factors that influence the diagnosis of autistic spectrum disorders. Soc Psychiatry Psychiatr Epidemiol 46, 1283–93 (2011).

39. D’Angelo, D. et al. Defining the Effect of the 16p11.2 Duplication on Cognition, Behavior, and Medical Comorbidities. JAMA Psychiatry 73, 20–30 (2016).

40. Kong, A. et al. Rate of de novo mutations and the importance of father’s age to disease risk. Nature 488, 471–5 (2012).

41. Fulco, C.J., Henry, K.L., Rickard, K.M. & Yuma, P.J. Time-Varying Outcomes Associated With Maternal Age at First Birth. Journal of Child and Family Studies 29, 1537–1547 (2020).

42. Lyall, K. et al. The Association Between Parental Age and Autism-Related Outcomes in Children at High Familial Risk for Autism. Autism Res 13, 998–1010 (2020).

43. Malaspina, D. et al. Paternal age and intelligence: implications for age-related genomic changes in male germ cells. Psychiatr Genet 15, 117–25 (2005).

44. Sanders, S.J. et al. A framework for the investigation of rare genetic disorders in neuropsychiatry. Nat Med 25, 1477–1487 (2019).

45. Brandler, W.M. et al. Paternally inherited cis-regulatory structural variants are associated with autism. Science 360, 327–331 (2018).

46. Lam, M. et al. RICOPILI: Rapid Imputation for COnsortias PIpeLIne. Bioinformatics 36, 930–933 (2020).

47. Lee, J.J. et al. Gene discovery and polygenic prediction from a genome-wide association study of educational attainment in 1.1 million individuals. Nat Genet 50, 1112–1121 (2018).

48. Lian, A., Guevara, J., Xia, K. & Sebat, J. Customized de novo mutation detection for any variant calling pipeline: SynthDNM. bioRxiv, 2021.02.10.427198 (2021).

49. Antaki, D., Brandler, W.M. & Sebat, J. SV2: Accurate Structural Variation Genotyping and De Novo Mutation Detection from Whole Genomes. Bioinformatics (2017).

50. Leppert, B. et al. A cross-disorder PRS-pheWAS of 5 major psychiatric disorders in UK Biobank. PLoS Genet 16, e1008185 (2020).

51. Chang, Y.S. et al. Reciprocal white matter alterations due to 16p11.2 chromosomal deletions versus duplications. Hum Brain Mapp 37, 2833–48 (2016).

52. Qiu, Y. et al. Oligogenic Effects of 16p11.2 Copy-Number Variation on Craniofacial Development. Cell Rep 28, 3320–3328 e4 (2019).

53. Qureshi, A.Y. et al. Opposing brain differences in 16p11.2 deletion and duplication carriers. J Neurosci 34, 11199–211 (2014).

54. Brunetti-Pierri, N. et al. Recurrent reciprocal 1q21.1 deletions and duplications associated with microcephaly or macrocephaly and developmental and behavioral abnormalities. Nat Genet 40, 1466–71 (2008).

55. Crespi, B.J. Autism As a Disorder of High Intelligence. Front Neurosci 10, 300 (2016).

